# Prevalence of SARS-CoV-2 among workers returning to Bihar gives snapshot of COVID across India

**DOI:** 10.1101/2020.06.26.20138545

**Authors:** Anup Malani, Manoj Mohanan, Chanchal Kumar, Jake Kramer, Vaidehi Tandel

## Abstract

India has reported the fourth highest number of confirmed SARS-CoV-2 cases worldwide. Because there is little community testing for COVID, this case count is likely an underestimate. When India partially exited from lockdown on May 4, 2020, millions of daily laborers left cities for their rural family homes. RNA testing on a near-random sample of laborers returning to the state of Bihar is used to estimate positive testing rate for COVID across India for a 6-week period immediately following the initial lifting of India’s lockdown. Positive testing rates among returning laborers are only moderately correlated with, and 21% higher than, Indian states’ official reports, which are not based on random sampling. Higher prevalence among returning laborers may also reflect greater COVID spread in crowded poor communities such as slums.

## Main text

The burden of SARS-CoV-2 (COVID) in India is high and rising. India has roughly the same population (1.378 billion (*1*)) as China, but already quadruple the number of cases. Since it is the world’s 2nd most populous country, it is unsurprising that India ranks 4th in number of cases and 8th in number of deaths reported as of 19 June 2020. Yet these absolute numbers likely drastically undercount the burden: India ranks 138th in the world in testing rate, at 4,659 tests per million (*2*).

There are no reliable and consistent estimates of the extent of the prevalence of infection across India. While India conducts testing in every state and territory, it is just beginning representative community testing (*3*). Early efforts by the Indian Council of Medical Research (ICMR) conflated prevalence among patients admitted for non-COVID symptoms with community prevalence (*4*). Moreover, most ongoing efforts to conduct community-level testing in India are serological, so they measure the share of population previously infected or recovered, rather than currently infected (*5*).

The number of cases reported on a daily basis by the Indian government comes from non-representative RNA testing. These data are products of testing policies and rates that vary across states and are hard to track, making interstate comparisons difficult (*6*). Some states have implemented tight restrictions on which patients can be tested, doctors’ ability to prescribe tests, and the number of labs that are permitted to conduct testing (*7, 8*).

Here, we rely on a unique dataset of RNA tests conducted on 43,356 workers from around the country who returned to their home state of Bihar between May 4 - June 10, 2020, to provide a snapshot of COVID burden across India. In the aftermath of the lockdown and ensuing job losses, India experienced a large flight of daily laborers (hereafter workers) migrating from urban areas back to their familial home states (*9*). Bihar (population 100 million) received roughly 2.1 million such workers between May 4 and June 10 (Supplement table 1). Bihar implemented a testing program that included a random sampling of returning workers. Combining data from this testing with demographic information on the returning workers, we present information on the prevalence of COVID in states across India, compare these positive test rates with those reported by the states, and report positive test rates by demographic group.

## Background

India’s first confirmed COVID case, on January 29, 2020, was a student who had returned home to Kerala state from Wuhan, China. Other states had introductions by foreign travelers who had visited other countries. The first recorded introduction in Bihar was on March 20, 2020, by a traveler from Qatar. After imposing a series of increasingly strict travel restrictions, India imposed a nation-wide lockdown on March 24, 2020. It was one of the most stringent lockdowns worldwide: mobility to workplaces and retail fell 48% and 69%, respectively (*10*).

India has nearly 11 million persons who work in one state, but regularly return to their rural homes in another (*11*). The lockdown was imposed 4 hours after it was announced and barred travel between districts, so many workers were unable to return to their rural homes. Many wanted to return home, as the lockdown prohibited work opportunities, especially in cities, which had a higher burden of COVID cases.

On May 4, India announced a partial release from lockdown, allowing workers to return to their out-of-state homes with the consent of destination states (*12*). Reverse migration surged to over 10 million persons within a few weeks (*9*). Roughly 2.8 million workers were expected to return to Bihar (*13*).

## Methods

We study the rate at which workers returning to Bihar tested positive for COVID during a 38 day period from May 4-June 10, 2020. Between May 4 and 21 (“period 1”), Bihar imposed a 14-day quarantine in government facilities on all returning workers. The government’s policy was to test all symptomatic workers, and randomly test the remaining workers, with a heavier weight on pregnant women, children under age 10, and elderly above 65 (*14*).

Between May 22 and 31 (“period 2”), Bihar focused its institutional quarantine policy on workers from a few cities that were believed to have higher rates of infections: the National Capital Region (NCR) around New Delhi, Mumbai and Pune in Maharashtra, Surat and Ahmedabad in Gujarat, Kolkata in West Bengal, and Bengaluru in Karnataka. For these workers, the testing policy remained the same as in period 1. All remaining workers were placed under 14-day home quarantine. They, or the members of their household, were tested only if they were symptomatic.

From June 1 to 10 (“period 3”), Bihar continued its quarantine policy from period 2. It tested workers in government quarantine as in period 1. For workers in home quarantine, Bihar switched to random testing of workers regardless of symptoms or demographics.

Returning workers were typically tested within 2 days of arriving in the state (Supplementary Figure 1). Initially, screening tests were conducted using Truenat machines; positive samples were confirmed by RT-PCR (*14*). The sensitivity and specificity of Truenat machines are 100% according to the manufacturer (*15*). After that was confirmed in a validation test on May 18, 2020, (*16*), the Indian Council of Medical Research stopped requiring RT-PCR confirmation (*17*). Therefore, we assume that the positive test rate measures COVID prevalence.

The data have information on the workers’ state of origin, age, and sex. Test results are aggregated from daily data up to roughly 2 week intervals, because there is a great deal of daily variation in the states from which workers arrive. The exact intervals correspond to periods during which testing policy is constant. For between-state comparisons, we drop states with less than 25 tests due to lack of precision.

To address concerns that the testing rate may indirectly be a function of disease status, i.e., selection bias, or that sampling weights are unknown, we do two things. First, we focus on subsamples, e.g., adult males, who were both selected at random and weighted equally in selection (Supplement Table 5A). Second, we estimate nonparametric bounds on prevalence, a method known as partial identification (*18*) (Supplement Table 5B). Lower and upper bounds assume all non-tested persons are uninfected and infected, respectively. Such upper bounds are uninformative because testing rates are so low (*19*). Therefore, we only present lower bounds.

## Results

### Testing rate

Of the 1.6 million workers who returned to Bihar from the origin states that met our inclusion criteria during all 3 periods in our study (Table 1), 43,356 were tested (Supplement Table 1), yielding an overall testing rate of 2.7%. This is higher than the average official testing rate of 0.1%, defined as the number of tests conducted by an origin state on their residents divided by that state’s population (Supplement Table 1). The number of tests conducted on returning workers was fairly constant at 1,141 per day. The ostensible rate at which workers were tested declined in period 2 and jumped in period 3, mainly because the number of workers per day increased in period 2 and then collapsed in period 3. Moreover, period 1 is nearly twice as long (18 v. 10 days) as later periods.

**Table 1.** Number of returning workers and testing rate, by state or territory of origin and date.

| State | (1) May 4-May 21 |  | (2) May 22-May 31 |  | (3) June 1-June 10 |  |
| --- | --- | --- | --- | --- | --- | --- |
|  | Returning Workers | Percent Tested | Returning Workers | Percent Tested | Returning Workers | Percent Tested |
| Andhra Pradesh | 17,161 | 2.6% | 10,894 | 1.7% | - | - |
| Chandigarh | 9,180 | 1.9% | 4,384 | 2.2% | - | - |
| Chhattisgarh | - | - | - | - | - | - |
| Delhi | 56,442 | 4.0% | 109,949 | 1.4% | 8,930 | 28.9% |
| Gujarat | 116,560 | 3.0% | 223,183 | 1.0% | - | - |
| Haryana | 51,733 | 2.8% | 50,420 | 1.5% | 2,609 | 40.9% |
| Jammu And Kashmir | - | - | 5,050 | 0.9% | - | - |
| Jharkhand | - | - | - | - | - | - |
| Karnataka | 46,406 | 1.5% | 54,386 | 0.5% | 16,361 | 3.0% |
| Madhya Pradesh | 2,982 | 11.4% | 2,774 | 5.1% | - | - |
| Maharashtra | 101,146 | 3.1% | 177,514 | 1.0% | 3,008 | 82.5% |
| Odisha | - | - | 1,753 | 4.9% | - | - |
| Punjab | 75,032 | 1.5% | 82,143 | 0.8% | 7,099 | 11.5% |
| Rajasthan | 40,691 | 3.8% | 42,183 | 1.2% | 1,600 | 27.1% |
| Tamil Nadu | 31,227 | 1.0% | 41,903 | 0.7% | 22,997 | 2.9% |
| Telangana | 36,464 | 3.4% | 18,636 | 1.3% | - | - |
| Uttar Pradesh | 64,813 | 2.4% | 50,812 | 1.1% | 1,200 | 49.9% |
| Uttarakhand | 4,314 | 1.1% | 4,328 | 1.8% | 4,754 | 1.1% |
| West Bengal | - | - | - | - | 3,875 | 10.9% |
| Total | 654,151 | 3.0% | 880,312 | 1.1% | 72,433 | 17.9% |

*Notes. Only states and territories from which at least 25 workers were tested each period are included in this table. States and periods for which data on the number of workers are missing are marked as such. Testing rate equals number of workers tested (from Supplement Table 1) divided by number of returning workers that period. Data on returning workers is based on information collected by the Government of Bihar on the number of people returning to the state by special passenger trains. It does not contain data on states neighbouring Bihar since returning workers used other means of travel to enter the state.*

### Prevalence among returning workers

Prevalence of COVID was 6.3% among returning workers in period 1 (Table 2). This rate rose (p<0.01) to 11% in period 2 and then declined (p<0.01) to 8.8% in period 3. Overall, the rate increased 40% during the duration of the study.

**Table 2.**
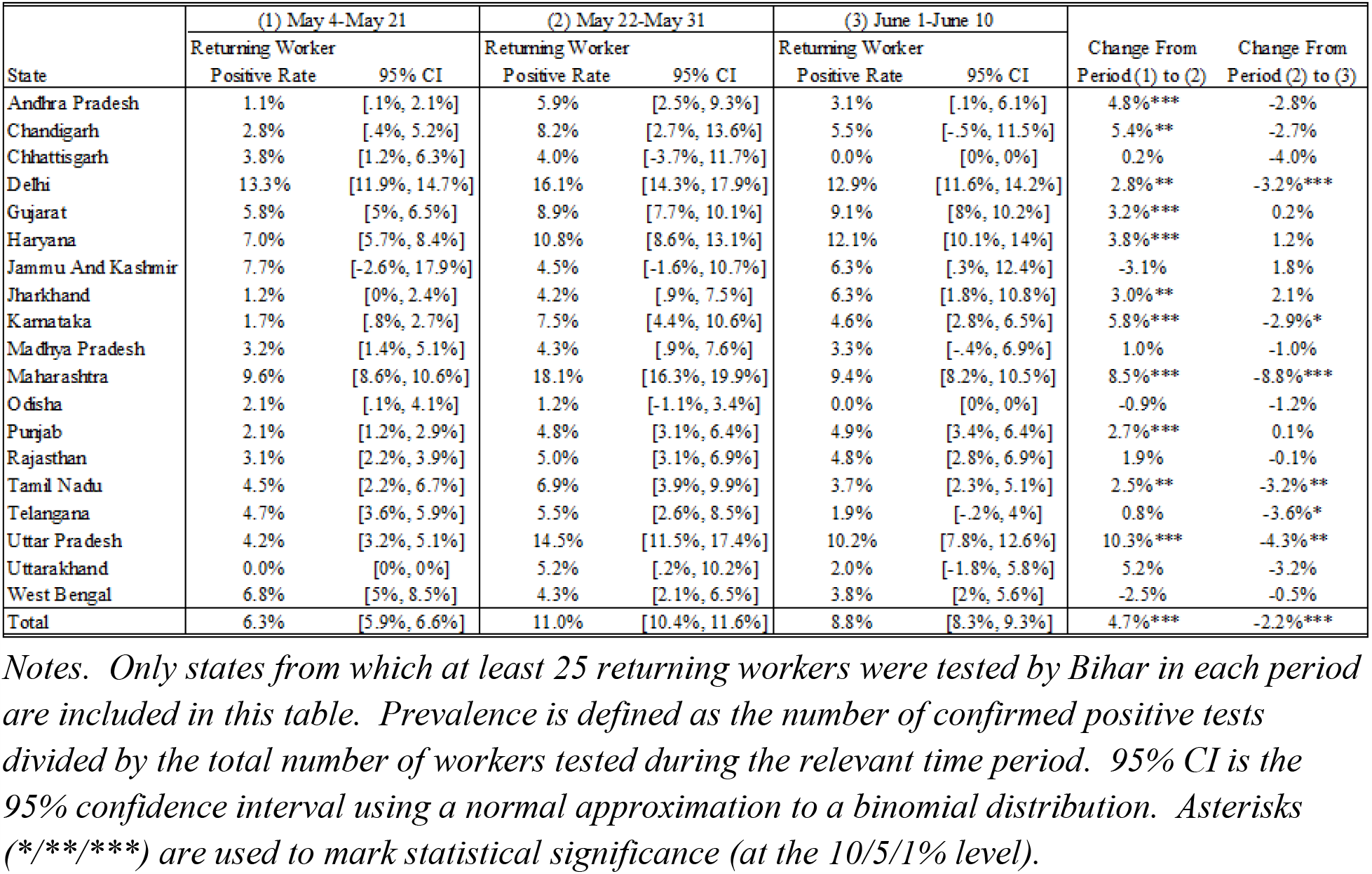
Prevalence among returning workers, by state or territory of origin and period.

These overall changes mask substantial variation in changes across states. In period 1, workers returning from Delhi (13.3%), Maharashtra (9.6%), and West Bengal (6.8%), had the highest prevalence (Table 2). Since these workers were largely employed in cities, these numbers likely reflect prevalence among the poor in urban centers such as New Delhi, Mumbai, Pune and Kolkata.

In period 2, prevalence remained high in workers from Delhi (16.1%) and Maharashtra (18.1%), though the latter now had the highest rate. However, Haryana (10.8%) and Uttar Pradesh (14.5%) had higher rates than West Bengal (4.3%) in this period. Growth was largest in Uttar Pradesh (+10.3%) and Maharashtra (+8.5%), while rates fell most in West Bengal (−2.5%) and Jammu and Kashmir (−3.1%). Notwithstanding these changes, the within-state correlation between the positive rate in the two periods is 0.66 (p<.01).

In period 3, prevalence was highest in workers from Delhi (12.9%), Haryana (12.1%) and Uttar Pradesh (10.2%). Workers from most states experienced declines in prevalence from period 2, with the largest, significant declines associated with Maharashtra (−8.8%), Uttar Pradesh (4.3%) and Delhi (−3.2%). The correlation between prevalence in periods 2 and 3 is high (*ρ*= 0.82 p<.01).

### Comparison with state testing results

The positive test rate observed among returning workers is moderately correlated with the positive test rates measured by workers’ origin states (*ρ*= 0.61, p<0.01). However, the positive rate among workers returning to Bihar was on average 1 percentage point (pp) (21%) higher than the prevalence reported by the origin state’s testing data across the 3 periods. Moreover, there is variation in the discrepancy between these two measures of COVID burden. Part of the variation is temporal. While positive rates rose gradually in the data from states, it rose and then fell among returning workers. Part of the variation is geographic. The discrepancy is widest in Maharashtra (6.7 pp), Haryana (5.5 pp), and Uttar Pradesh (4.7 pp) and lowest in Delhi (0 pp), Madhya Pradesh (0.5 pp), and Odisha (0.8 pp).

### Selection bias

A challenge with making state-wise comparisons using official data is that states adopted varying testing policies with large differences in testing rates (Supplement Table 1) and composition of individuals tested. This is somewhat mitigated when using data from returning workers because all testing was done under a common testing policy by Bihar.

However, even the representativeness of Bihar’s testing may be questioned because the state had different sampling weights for symptomatic persons; pregnant women, children and the elderly; and remaining persons. Moreover, the state did not estimate the number of migrants in each health and demographic bin, so sampling weights cannot be computed.

To address sampling bias, Supplement Table 5A presents the prevalence for different groups of workers (symptomatic workers, non-symptomatic workers, non-child and non-elderly men). The correlation between prevalence in these subgroups and overall prevalence among workers was quite high (versus prevalence for whole sample all periods: *ρ*= 0.36, p<0.01 for symptomatic; *ρ*= 0.36, p<0.01 for non-symptomatic; *ρ*= 0.47, p<0.01 for men ages 10 to 65).

The Supplement also presents a lower bound on the positive rate for both data from origin states and from workers returning to Bihar (Supplement Table 5B). The lower bounds for Delhi and Maharashtra workers remain the highest among states across all periods. However, the correlation between lower bounds based on states’ own testing and tests of returning workers across all 3 periods is weak (*ρ*= 0.31, p=0.05).

### Prevalence by demographic group

Prevalence was lowest among girls under 10 (Table 4), who had significantly lower rates than any group other than adult women (Supplement Table 7). Prevalence was highest among elderly and non-elderly men, though only the differences between these two groups and children (of both sexes) were significant.

**Table 3.** Difference between positive test rates among returning workers and among residents of state, by state or territory of origin and period.

| State | (1) May 4-May 21 |  | (2) May 22-May 31 |  | (3) June 1-June 10 |  |
| --- | --- | --- | --- | --- | --- | --- |
|  | State-Reported Positive Rate | Difference | State-Reported Positive Rate | Difference | State-Reported Positive Rate | Difference |
| Andhra Pradesh | 0.5% | 0.6%* | 0.6% | 5.3%*** | 0.9% | 2.2%*** |
| Chandigarh | 6.7% | 3.9%** | 5.9% | 2.3% | 4.7% | 0.7% |
| Chhattisgarh | 0.3% | 3.5%*** | 1.5% | 2.5% | 3.0% | 3.0% |
| Delhi | 7.5% | 5.8%*** | 14.3% | 1.8%** | 24.6% | 11.8%*** |
| Gujarat | 8.7% | 3.0%*** | 8.9% | 0.0% | 8.6% | 0.5% |
| Haryana | 1.0% | 6.0%*** | 3.8% | 7.0%*** | 8.4% | 3.7%*** |
| Jammu And Kashmir | 0.9% | 6.8%*** | 1.7% | 2.8% | 2.9% | 3.4% |
| Jharkhand | 0.7% | 0.5% | 1.3% | 2.9%*** | 2.9% | 3.4%** |
| Karnataka | 1.0% | 0.7%* | 1.4% | 6.1%*** | 2.5% | 2.1%*** |
| Madhya Pradesh | 4.1% | 0.9% | 5.0% | 0.7% | 3.2% | 0.1% |
| Maharashtra | 17.4% | 7.8%*** | 18.1% | 0.0% | 18.8% | 9.5%*** |
| Odisha | 1.4% | 0.7% | 2.0% | 0.8% | 3.9% | 3.9%** |
| Punjab | 2.6% | 0.5% | 0.9% | 3.9%*** | 0.9% | 4.0%*** |
| Rajasthan | 2.2% | 0.9%** | 1.9% | 3.1%*** | 2.1% | 2.7%*** |
| Tamil Nadu | 5.0% | 0.5% | 7.1% | 0.2% | 9.8% | 6.1%*** |
| Telangana | - | - | - | - | - | - |
| Uttar Pradesh | 2.5% | 1.6%*** | 3.1% | 11.4%*** | 3.0% | 7.1%*** |
| Uttarakhand | 0.8% | 0.8% | 5.2% | 0.0% | 6.9% | 4.9% |
| West Bengal | 2.2% | 4.6%*** | 2.6% | 1.7%* | 4.2% | 0.4% |
| Total | 4.5% | 1.8%*** | 5.5% | 5.6%*** | 6.4% | 2.4%*** |

Notes. Statistics for states from which testing results are not available are marked as missing. For some states, the dates for test result data do not correspond exactly to the dates of each of the 3 periods; in those cases, we take data for the closest period corresponding to each of the 3 periods. State-reported positive rate is the number of confirmed cases reported by a state divided by the number of tests conducted by that state during the relevant time period. Asterisks (\*/\*\*/\*\*\*) are used to mark statistical significance (at the 10/5/1% level).

**Table 4.**
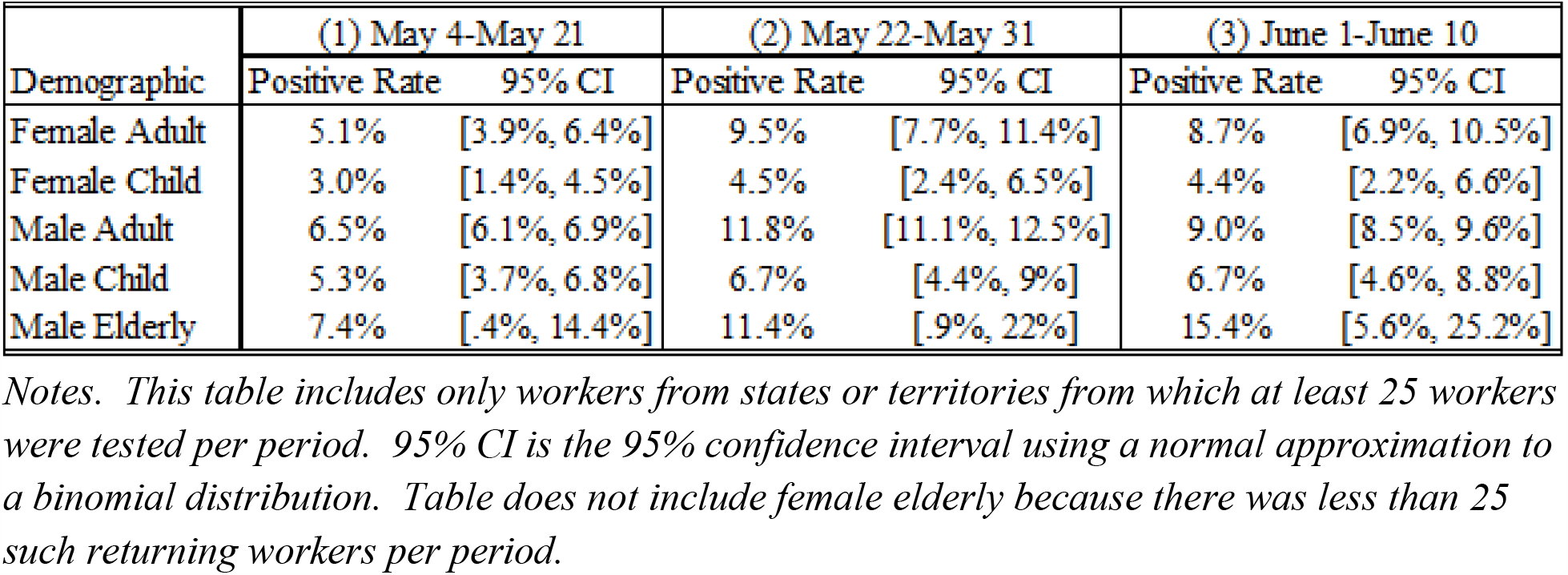
Prevalence among returning workers, by demographic group.

| Demographic | (1) May 4-May 21 |  | (2) May 22-May 31 |  | (3) June 1-June 10 |  |
| --- | --- | --- | --- | --- | --- | --- |
|  | Positive Rate | 95% CI | Positive Rate | 95% CI | Positive Rate | 95% CI |
| Female Adult | 5.1% | [3.9%, 6.4%] | 9.5% | [7.7%, 11.4%] | 8.7% | [6.9%, 10.5%] |
| Female Child | 3.0% | [1.4%, 4.5%] | 4.5% | [2.4%, 6.5%] | 4.4% | [2.2%, 6.6%] |
| Male Adult | 6.5% | [6.1%, 6.9%] | 11.8% | [11.1%, 12.5%] | 9.0% | [8.5%, 9.6%] |
| Male Child | 5.3% | [3.7%, 6.8%] | 6.7% | [4.4%, 9%] | 6.7% | [4.6%, 8.8%] |
| Male Elderly | 7.4% | [.4%, 14.4%] | 11.4% | [.9%, 22%] | 15.4% | [5.6%, 25.2%] |

Notes. This table includes only workers from states or territories from which at least 25 workers were tested per period. 95% CI is the 95% confidence interval using a normal approximation to a binomial distribution. Table does not include female elderly because there was less than 25 such returning workers per period.

## Discussion

### Comparison with states’ data

While prevalence among workers returning to Bihar was moderately correlated with the positive test rate reported by origin states, there are important differences. One is that, while the positive test rate rose gradually in state reports, it rose and then fell in data on returning workers. It is possible that changes in Bihar’s testing policy or the composition of returning workers explains the discrepancy.

However, the non-uniform changes in prevalence among workers from different states suggest that Bihar’s testing policy, which is largely uniform, is not responsible. Moreover, the variable changes in prevalence over time among workers across states suggest that selection of workers is not the explanation. An alternative explanation for the discrepancy is that data on returning workers may be more representative than state data of the non-monotonic patterns in prevalence common in epidemics. A national epidemic is a sum of local epidemics (*20*). The data on workers is compatible with this view.

### External validity

Returning workers may not be representative of the entire state or city population. These workers, often daily laborers, tend to be less wealthy as measured by consumption (cite). They likely live in more dense communities (*11*), which may increase COVID transmission.

However, workers in our sample are likely to be representative of poor communities in their origin cities. Testing was unlikely to detect infections that the workers acquired on the crowded trains used to transport workers home (*9*). Train journeys were less than 1 day and Bihar tested most returning workers within 2 days of arrival. RT-PCR does not yield positive results within 2 days of infection (*21*).

### Imperfect sensitivity

Given Indian Council of Medical Research’s validation of Bihar’s testing procedure, we equate positive test results with prevalence. However, imperfect specimen collection could lead to lower sensitivity than rates estimated in a lab setting (*22*), which would lead to an underestimation of infection rates Unfortunately, it is difficult to estimate the impact of imperfect specimen collection on sensitivity. If it were possible, one would adjust prevalence upwards using standard formulas (*23*).

### Policy implications

The analysis here suggests that random testing of travellers may be a useful method of national surveillance when there is variability in local testing protocols and rates. Moreover, to ensure prevalence is correctly estimated, backchecks should be conducted by experienced technicians to determine if sensitivity of testing in the field corresponds to that in the lab. Finally, our results suggest that poor communities in India may have higher prevalence of COVID than suggested by official statistics, perhaps due to crowding in their densely populated communities. These communities may require extra attention and support in infection control policies.

## Data Availability

Data may be requested from the Government of Bihar.

## Notes

### Competing Interest Statement

The authors have declared no competing interest.

### Funding Statement

This paper has no outside funding. Jake Kramer is funded by an internal grant from the University of Chicago's Center for Global Health.

### Author Declarations

The study has been reviewed by University of Chicago's SBS IRB (IRB20-1110) and has been found exempt.

## References

1. United Nations Department of Economic and Social Affairs Population Division, “2019 Revision of World Population Prospects,” (2019).

2. Worldometer, “COVID-19 Coronavirus Pandemic,” (2020).

3. Special Correspondent, “ICMR to test for community transmission in 75 districts,” The Hindu (10 May 2020).

4. N. Gupta et al., Severe acute respiratory illness surveillance for coronavirus disease 2019, India, 2020. Indian Journal of Medical Research 151, 236–240 (2020).

5. T. Thacker, “ICMR to check for community spread,” The Economic Times (8 May 2020).

6. S. Alexander, S. Devulapalli, “Everything you wanted to know about India’s test numbers, in five charts,” LiveMint (10 May 2020).

7. R. Chakraborty, in Hindustan Times. (2020).

8. A. Saikia, “Testing is key to fighting coronavirus – so why does India have such a low testing rate?,” Scroll.in (27 March 2020).

9. R. Roy, V. Agarwal, “Millions of Indians Are Fleeing Cities, Raising Fears of a Coronavirus ‘Land Mine’ in Villages,” Wall Street Journal (27 May 2020).

10. Google, “COVID-19 Community Mobility Report - India 29 May 2020 - Mobility changes,” (2020).

11. C. Imbert, “Covid-19: Expected migrant movement as lockdown eases,” Ideas for India (2020).

12. Government of India, “Order No. 40-3/2020/DM-I(A) (May 1, 2020),” (2020).

13. M. I. Khan, “First Batch of Stranded Migrant Workers Arrive in Bihar on Special Train,” News Click (2 May 2020).

14. Directions for Random Testing (2020 https://prsindia.org/files/covid19/notifications/4896.BR_CM_Directions_Random_Testing_May_7.pdf).

15. Molbio Diagnostics Pvt. Ltd. (2020).

16. Performance evaluation of TruenatTM SARS CoV-2 test on TruelabTM workstation: Validation Report (18 May 2020).

17. Revised Guidelines for TrueNat testing for COVID-19 (May 19, 2020).

18. F. Molinari, Econometrics with partial identification. The Handbook of Economet, (2019).

19. C. F. Manski, F. Molinari, Estimating the COVID-19 infection rate: Anatomy of an inference problem. Journal of Econometrics, (2020).

20. M. Zheng et al., Multiple peaks patterns of epidemic spreading in multi-layer networks. Chaos, Solitons Fractals 107, 135–142 (2018).

21. A. Tahamtan, A. Ardebili, Real-time RT-PCR in COVID-19 detection: issues affecting the results. Expert Review of Molecular Diagnostics 20, 453–454 (2020).

22. P. Wikramaratna, R. S. Paton, M. Ghafari, J. Lourenco, Estimating false-negative detection rate of SARS-CoV-2 by RT-PCR. medRxiv, 2020.2004.2005.20053355 (2020).

23. W. J. Rogan, B. Gladen, Estimating prevalence from the results of a screening test. American journal of epidemiology 107, 71–76 (1978).

